# Forecasting COVID-19 cases in US states using reconstructed incidence data

**DOI:** 10.1101/2025.10.20.25338346

**Authors:** Rebecca K Nash, Sangeeta Bhatia, Jack Wardle, Anne Cori, Pierre Nouvellet

## Abstract

Branching process models are commonly used in infectious disease forecasting and often rely on daily incidence data, but their utility can be restricted if incidence is not reported daily or if reporting becomes less frequent during prolonged outbreaks. In this study, an Expectation Maximisation algorithm is used to reconstruct the daily incidence of COVID-19 cases from weekly case counts. Using data from 13 US states that maintained mostly daily reporting of COVID-19 cases from March 2020 to February 2022, we evaluate forecasting performance by comparing models using the true daily incidence with those using reconstructed daily incidence. Our results show that forecasts generated from reconstructed incidence perform equally well as those generated using true daily incidence. These findings demonstrate the viability of using reconstructed incidence data for real-time forecasting, which could be particularly useful in scenarios where maintaining daily reporting is unsustainable or in settings with limited surveillance capacity.

## Introduction

Infectious disease forecasts are a crucial element of outbreak response, allowing us to estimate the likely number of future cases, hospitalisations or deaths. These predictions allow public health agencies to implement timely interventions and allocate resources more effectively to mitigate the impact of disease spread.

One common type of forecasting model is the branching process model, which is often based on the renewal equation and relies on past daily incidence data, the latest estimate for the time-varying reproduction number (*R*_*t*_ - the average number of cases caused by a primary case infected at time *t* of an outbreak, assuming that conditions remain the same after time *t*) and either the serial interval (SI) or generation time. Using these inputs, future incidence can be projected over a specified time window assuming that *R*_*t*_ remains constant. Throughout the COVID-19 pandemic, caused by the SARS-CoV-2 virus, branching process models have been used to forecast future cases, hospitalisations and deaths to inform real-time response globally [1–3].

In practice, the incidence of many infectious diseases is not reported daily and may instead be reported irregularly or as weekly counts, which can be a barrier to the use of these models in such contexts [4]. For example, due to the protracted nature of the COVID-19 pandemic and the demand on resources, many countries, regions and public health agencies did not maintain daily reporting of COVID-19 incidence [5–8]. Although methods have been developed to reconstruct daily incidence from aggregated counts for the purpose of estimating transmissibility [9, 10], the impact of using reconstructed incidence on the accuracy of forecasts is unknown. Understanding these implications is important not only for determining the viability of using reconstructed incidence for forecasting when daily incidence is unavailable, but also for informing the design of surveillance systems for future epidemics. To investigate this, we use the incidence of COVID-19 cases in the United States (US) as a case study.

In the US, the first laboratory-confirmed case of COVID-19 was reported in Washington state on 20th January 2020 following recent travel to Wuhan, China [11]. On 11th March 2020, the World Health Organisation officially declared COVID-19 a pandemic [12]. In the two years following this declaration (March 2020 - February 2022), the US experienced six distinct outbreak phases, which we have defined as the: ‘Early Pandemic’, ‘Summer Surge’, ‘Winter Surge’, ‘Vaccine Rollout’, ‘Delta Wave’ and ‘Omicron Wave’ (Fig 1).

**Fig 1.**
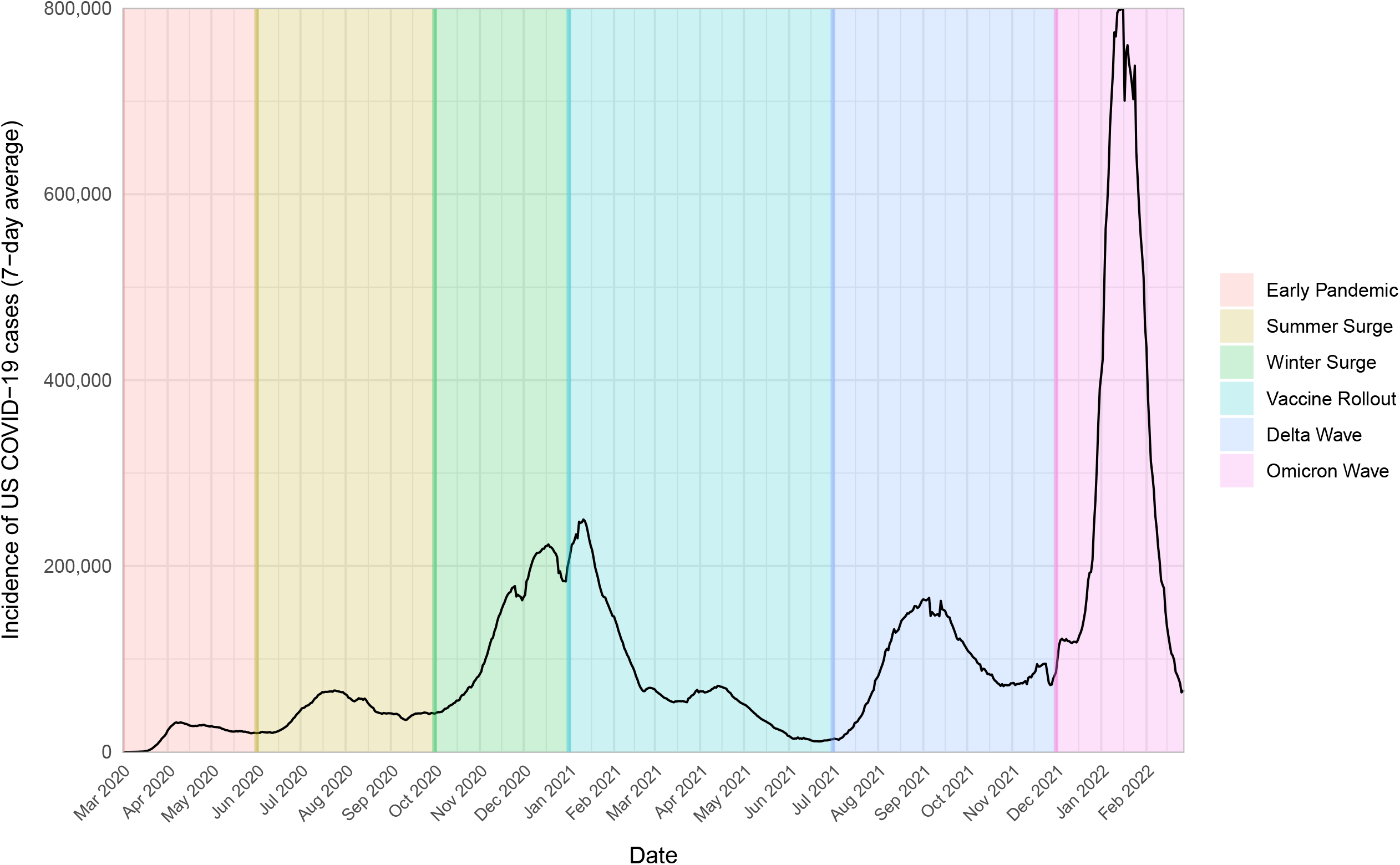
Incidence of COVID-19 cases in the US (7-day rolling average) from March 2020 to February 2022. Highlighted periods represent key phases of the pandemic, including: ‘Early Pandemic’ (March - May 2020), ‘Summer Surge’ (June - September 2020), ‘Winter Surge’ (October - December 2020), ‘Vaccine Rollout’ (January - June 2021), ‘Delta Wave’ (July - November 2021) and ‘Omicron Wave’ (December 2021 - February 2022). See text for a description of each phase.

During the ‘Early Pandemic’ (March - May 2020), containment measures such as social distancing, school and workplace closures and stay-at-home orders began to be implemented to curb the virus’s spread [13]. Each US state had the autonomy to impose their own countermeasures and each had their own approach to reporting incidence data [14]. Across states there was considerable variation in the interventions implemented, when they were implemented and their duration [15].

The ‘Summer Surge’ (June - September 2020) followed the relaxation of initial lockdowns, resulting in a rise in case numbers in many states. A more substantial ‘Winter Surge’ (October - December 2020) then coincided with public holiday gatherings and colder weather, which led to increased indoor activities and travel [16].

Cases began to steadily decline during the ‘Vaccine Rollout’ phase (January - June 2021) and many US states reduced the reporting frequency of cases and deaths, typically shifting to reporting 4-5 days per week [7, 17].

Despite the reduction in reporting frequency across many US states, two waves of COVID-19 cases occurred: the ‘Delta Wave’ (July - November 2021), thought to have been partly driven by uneven vaccination coverage [17], and the largest ‘Omicron Wave’ (December 2021 - February 2022), driven by the emergence of the highly transmissible, though less severe, Omicron variant [18].

From March 2022 onward, the policy approach to COVID-19 shifted toward endemicity across the US, focusing on long-term management strategies for COVID-19 [19]. At this stage, the vast majority of US states had ceased daily reporting of cases and deaths.

In this study, we aim to determine whether daily reconstructed data can reliably support accurate forecasting and response planning in instances where daily reporting has ceased. Additionally, we assess whether maintaining daily reporting offers a significant advantage over the use of reconstructed data. To this end, we use COVID-19 case incidence from 13 US states that maintained mostly daily reporting as a case study. We compare the forecasting performance of a branching process model using the original daily reported incidence and daily incidence reconstructed from a simulated weekly reporting scenario.

## Materials and methods

### COVID-19 data

Incidence of COVID-19 cases in the US were obtained from the COVID-19 Data Repository by the Center for Systems Science and Engineering at Johns Hopkins University [20, 21]. The time series chosen encompasses two years of COVID-19 circulation in the US from March 2020 to February 2022. Case incidence data for 13 states were chosen for the analysis as they maintained mostly daily reporting throughout this time period. These states include California, Colorado, Delaware, Hawaii, Maryland, Missouri, New Jersey, New York, North Dakota, Ohio, Pennsylvania, Tennessee and Texas (accounting for around 45% of the US population [22]). Day of the week reporting patterns of case incidence data by day of the week were analysed by state and outbreak phase (Fig 1) by calculating the percentage of weekly cases reported on each day. Daily percentages were summarised across weeks using boxplots, which display the median, interquartile range, and overall spread of percentages (excluding outliers).

### Expectation Maximisation algorithm

For each included US state, we artificially aggregated the reported daily incidence data to weekly counts and reconstructed daily incidence data using an Expectation Maximisation (EM) algorithm previously described by the study authors [9]. Briefly, the time-varying reproduction number, *R*_*t*_, is estimated for each aggregation window in turn, translated into a growth rate and used to iteratively reconstruct daily incidence. The method ensures that, if the reconstructed incidence were to be reaggregated, it would still sum to the original totals. The EM algorithm is implemented using the EpiEstim R package [23], and both the reported and reconstructed daily incidence data were supplied as inputs to the forecasting model.

### Model

The renewal equation (Eq (1)) is used to forecast the future incidence of COVID-19 cases [24]. The model assumes that the incidence of new cases on day *t* (*I*_*t*_) can be represented by a Poisson process:

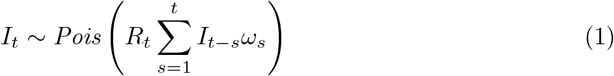

where *R*_*t*_ is the time-varying reproduction number and the past incidence (*I*_*t*−*s*_) is weighted by *ω*_*s*_, the probability mass function of the SI [25, 26]. The SI of COVID-19 is believed to have varied throughout the pandemic, becoming shorter with the emergence of new variants [27]. To provide a middle ground, the SI of mean 4.8 days and standard deviation 2.7 days was chosen for this analysis [28]. To calibrate the model, *R*_*t*_ is estimated using case incidence within a fixed calibration window prior to the projection date, where *R*_*t*_ is assumed to remain constant. To smooth out ‘weekend effects’ in the data [9] a 10-day calibration window was chosen, following the approach of Bhatia et al [29]. For the period before the calibration window, combinations of *R*_*t*_ and case incidence consistent with the observed incidence in the calibration window were jointly estimated using Markov Chain Monte Carlo (MCMC) implemented within the ‘jointlyr’ R package [24, 30]. The MCMC was run for 10,000 iterations, from which 1,000 sets of *R*_*t*_ and back-calculated incidence were sampled. For each sampled set, 10 stochastic realisations of the projected daily incidence of cases were drawn [29].The resulting 10,000 trajectories (1,000 parameter sets x 10 stochastic realisations) were summarised by computing quantiles (0.025, 0.25, 0.5, 0.75, 0.975) for each forecast day. See Bhatia et al for further details [29].

Using this model, 28-day forecasts were produced for each week of the dataset between 9th March 2020 and 27th February 2022. Projections were initiated on each day of the week to explore whether the starting day influenced forecast performance. The 28-day forecasts using reported and reconstructed incidence data were then aggregated to weeks (forecast weeks 1-4) and their ability to capture the true weekly cases in each forecast week was compared. Projection weeks for certain states were excluded if zero cases were reported in the previous week or if non-daily reporting was suspected (a conservative threshold was used: more than 30 cases reported in a week with at least one day of zero incidence) within the calibration window.

Forecasting performance was assessed using the Continuous Ranked Probability Score (CRPS) metric, which is a measure of the absolute difference between the observed weekly cases in the forecast period and the forecast distribution. Therefore, the lower the CRPS value, the closer the forecast is to the true observed cases for that forecast week. In order to mitigate the effect of the varying magnitude of case counts across the included US states, a transformation of *log*(*x* + 1) was applied to the true cases and the predictions, as recommended by Bosse et al [31]. The CRPS was then calculated using the ‘scoringutils’ R package, which for this metric acts as a wrapper for the ‘scoringRules’ R package [32, 33]. CRPS values were then transformed using the base 10 logarithm and compared between the included US states and the outbreak time periods as defined in Fig 1.

## Results

Hereafter we refer to reported and reconstructed incidence data - these are the reported daily incidence and the daily incidence that has been reconstructed from weekly aggregated data, respectively.

### Reporting patterns by US state

The relative reporting of US COVID-19 cases on each day of the week compared to the weekly total varied according to the US state (Fig 2). Aggregated across all included states, the median percentage of weekly reported cases was lower on Sundays and Mondays, with a peak midweek on Thursdays and Fridays. All 13 included states generally reported fewer cases on Sundays, Mondays or Tuesdays. Colorado and North Dakota displayed a typical ‘weekend-effect’ with lower reporting on weekends and high reporting at the start of the week. Most states, excluding Colorado, Delaware, North Dakota and Texas, had highest case counts on Thursdays or Fridays. The degree of week-to-week variability in daily reporting patterns differed between states. States such as Hawaii and Delaware showed more inconsistency in the percentage of weekly cases reported on different days of the week, whereas states such as Colorado and New York exhibited more consistent and stable reporting trends.

**Fig 2.**
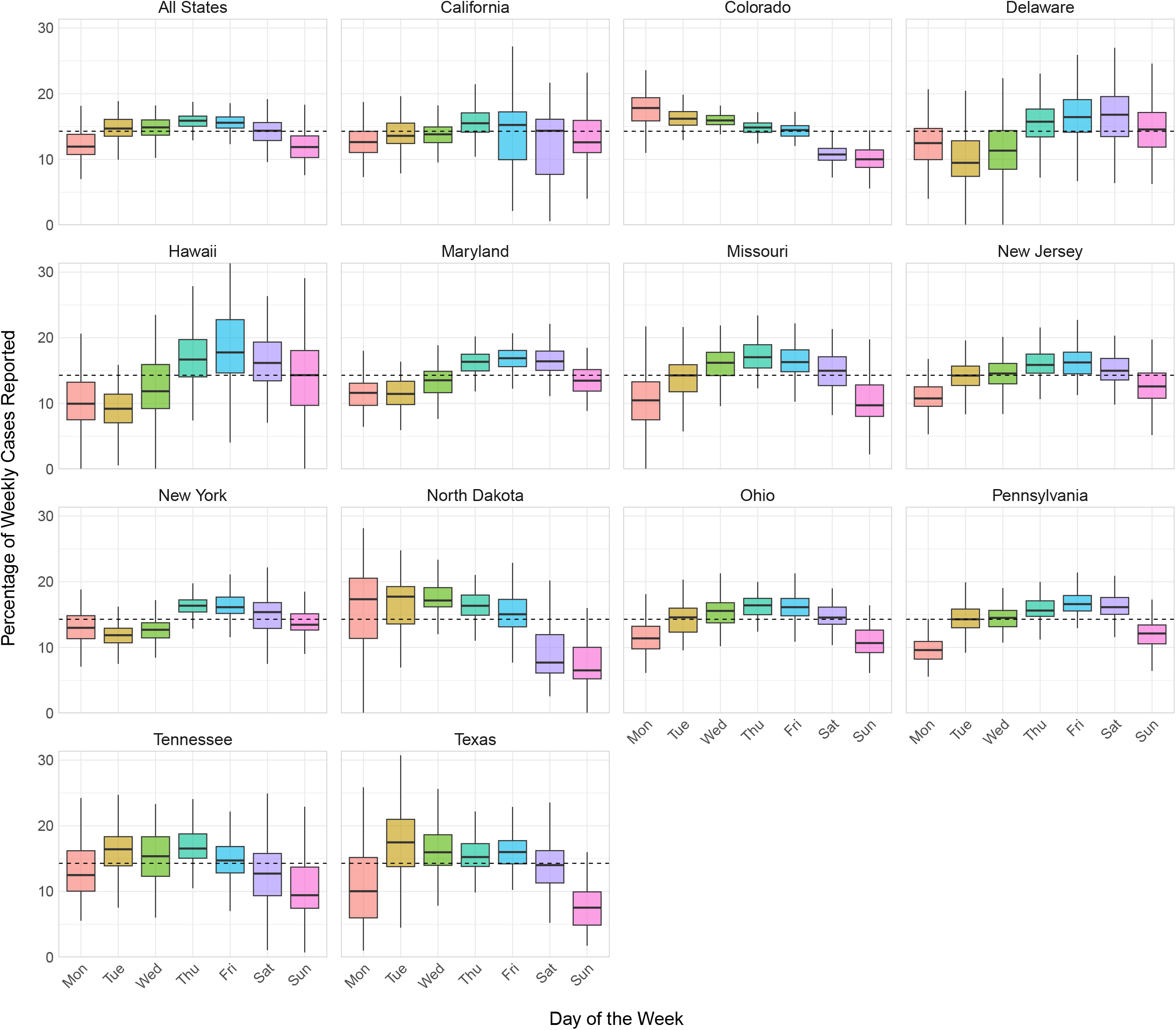
Relative reporting of COVID-19 cases by day of the week for each US state. Boxplots show the distribution of the percentage of weekly cases reported by day of the week, across weeks. Boxes represent the interquartile range (IQR), lines show medians, and whiskers extend to the most extreme values within 1.5 *×* IQR. Dashed lines represent the expected percentage if cases were reported equally on each day.

### Reporting patterns by outbreak phase

The day of the week pattern of COVID-19 case reporting also varies by outbreak phase (Fig 3). The Summer Surge, Winter Surge, Vaccine Rollout and Delta Wave phases exhibited a similar overall trend to that observed when aggregating across all phases, with a smaller median percentage of weekly cases reported on Sundays and Mondays, with peaks in reporting typically occurring midweek. The median percentage of cases reported peaked on Thursdays for the Vaccine Rollout and Delta Wave, while reported cases typically peaked on Tuesdays and Fridays for the Summer and Winter Surges, respectively. The Early Pandemic exhibited a different trend, with most cases reported from Thursdays to Saturdays, with lowest reporting on Tuesdays. The Omicron Wave, showed the most irregular pattern, with peaks on Thursdays and lowest reporting on Saturdays. Both the Delta Wave and Omicron Wave showed considerable week-to-week variability in daily reporting patterns, particularly on Sundays and Mondays.

**Fig 3.**
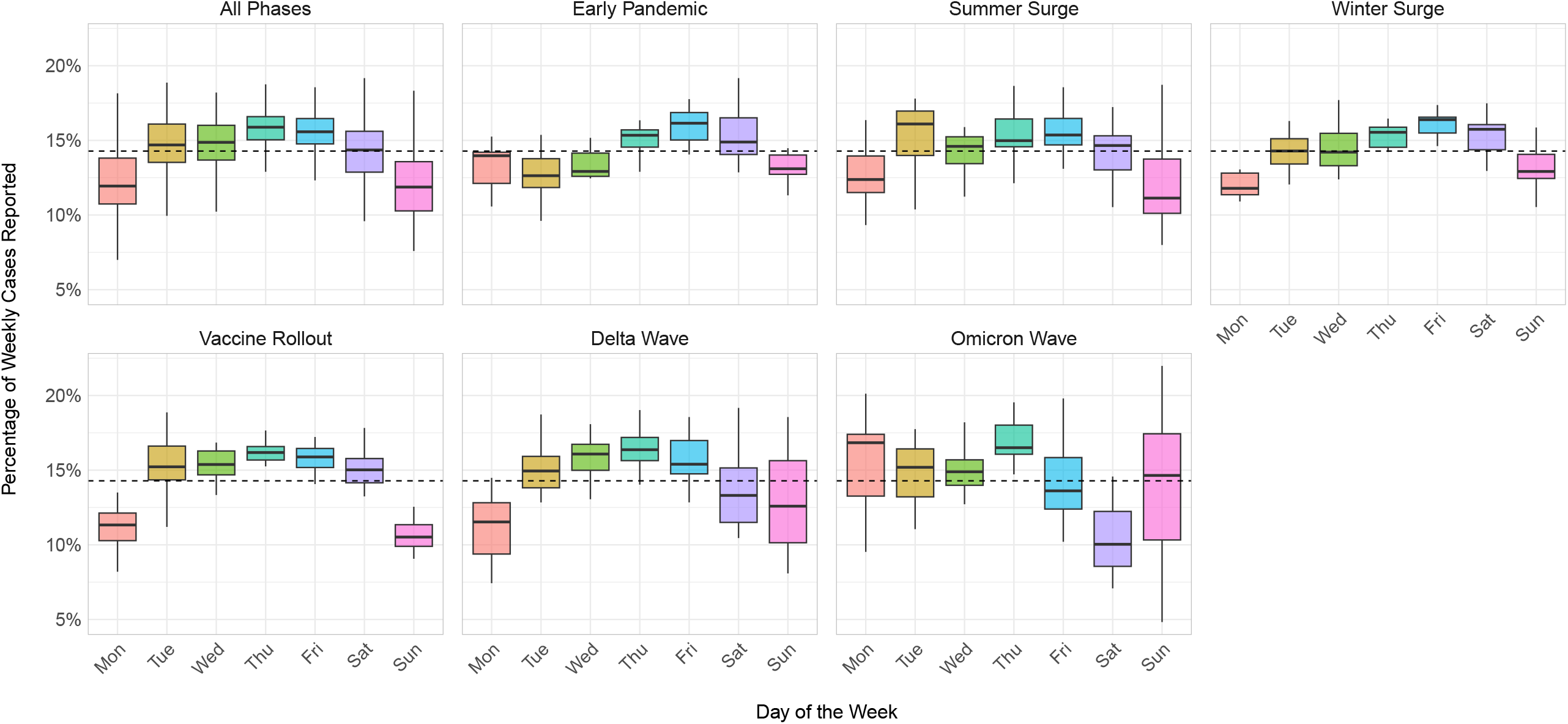
Relative reporting of COVID-19 cases by day of the week for each outbreak phase. Each phase corresponds to those shown in Fig 1. Boxplots show the distribution of the percentage of weekly cases reported by day of the week, across weeks. Boxes represent the interquartile range (IQR), lines show medians, and whiskers extend to the most extreme values within 1.5 *×* IQR. Dashed lines represent the expected percentage if cases were reported equally on each day.

### Forecasting performance by US state

Averaged across all 13 states included in the analysis, CRPS values showed only marginal differences within each forecast week (Fig 4), irrespective of whether reported or reconstructed incidence were used or the day of the week chosen for projections. As expected, CRPS values progressively increased over the forecast weeks, reflecting reduced performance over longer forecasting horizons.

**Fig 4.**
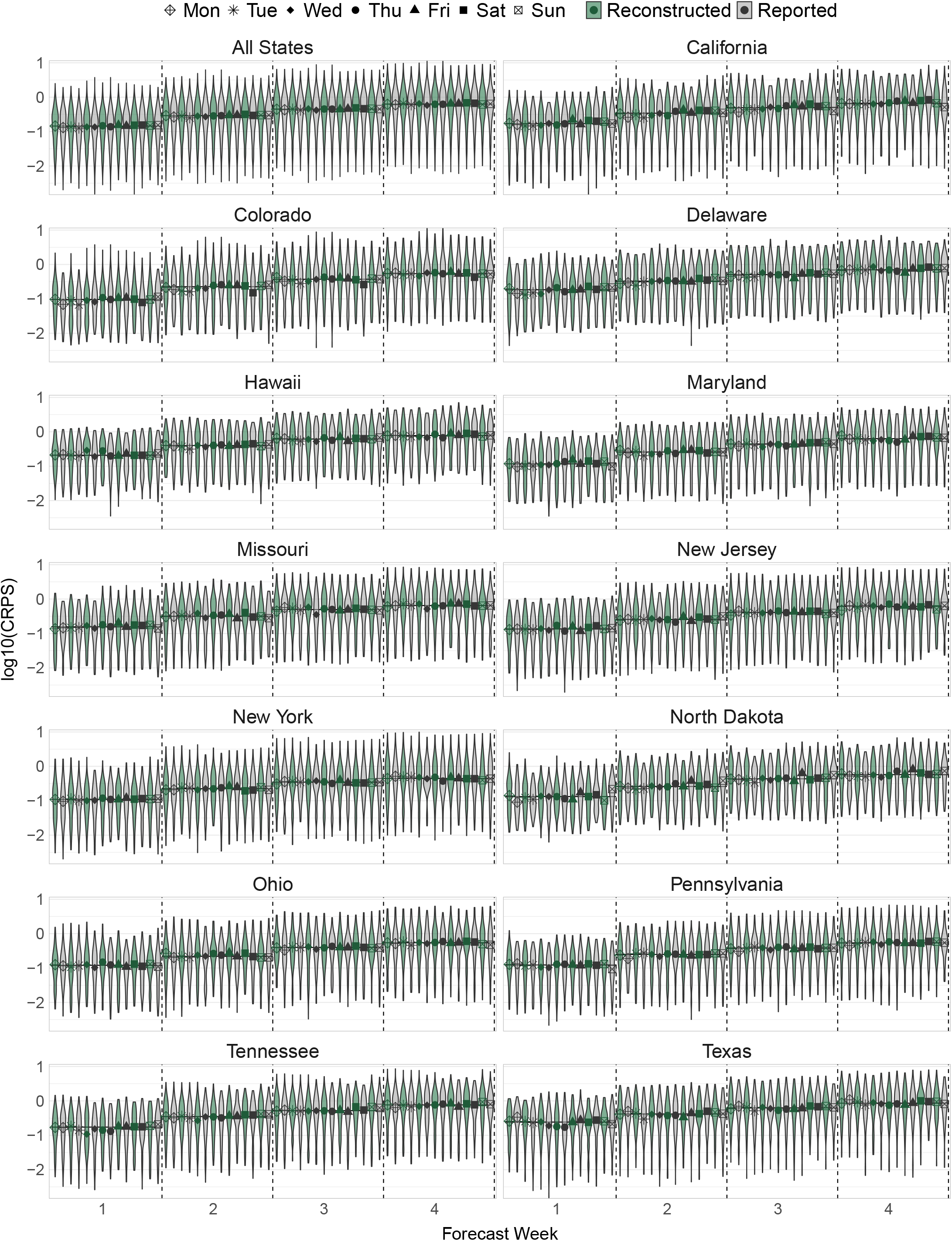
Violin plot showing the distribution of log-transformed (base 10) CRPS values by US state, based on the difference in true observed weekly cases and weekly forecasts produced from reconstructed (green) and reported (grey) incidence data. Points denote the median CRPS values, with symbols representing the projection day (last day of past incidence data supplied to the model): Mondays (diamonds with cross) Tuesdays (stars), Wednesdays (solid diamonds), Thursdays (circles), Fridays (triangles), Saturdays (solid squares) and Sundays (squares with cross). The solid black line represents the overall median CRPS score for the forecast week. The “All States” panel shows the distribution of CRPS values across all weeks and all individual state forecasts.

When considered separately, each state follows the same general trend of increasing CRPS over the forecast weeks (Fig 4). Within each forecast week, CRPS values are typically similar, but for some states there are small differences in performance depending on the projection day. For example, Friday and Sunday forecasts based on reported data appear to perform consistently worse (higher median CRPS values) in North Dakota.

### Forecasting performance by outbreak time period

When considering outbreak time periods separately (Fig 5), forecasting performance was worst overall (higher combined median CRPS in each forecast week) during the Early Pandemic (March - May 2020) and the large Omicron Wave (December 2021 - February 2022). During the Early Pandemic, forecasts using reported incidence data perform slightly better, other than Saturdays in the final forecast week. During the Omicron Wave, Wednesday forecasts using reported data perform best. In the Summer Surge (June - September 2020), Winter Surge (October - December 2020), Vaccine Rollout (January - June 2021) and Delta Wave (July - November 2021), differences between forecasts appear negligible.

**Fig 5.**
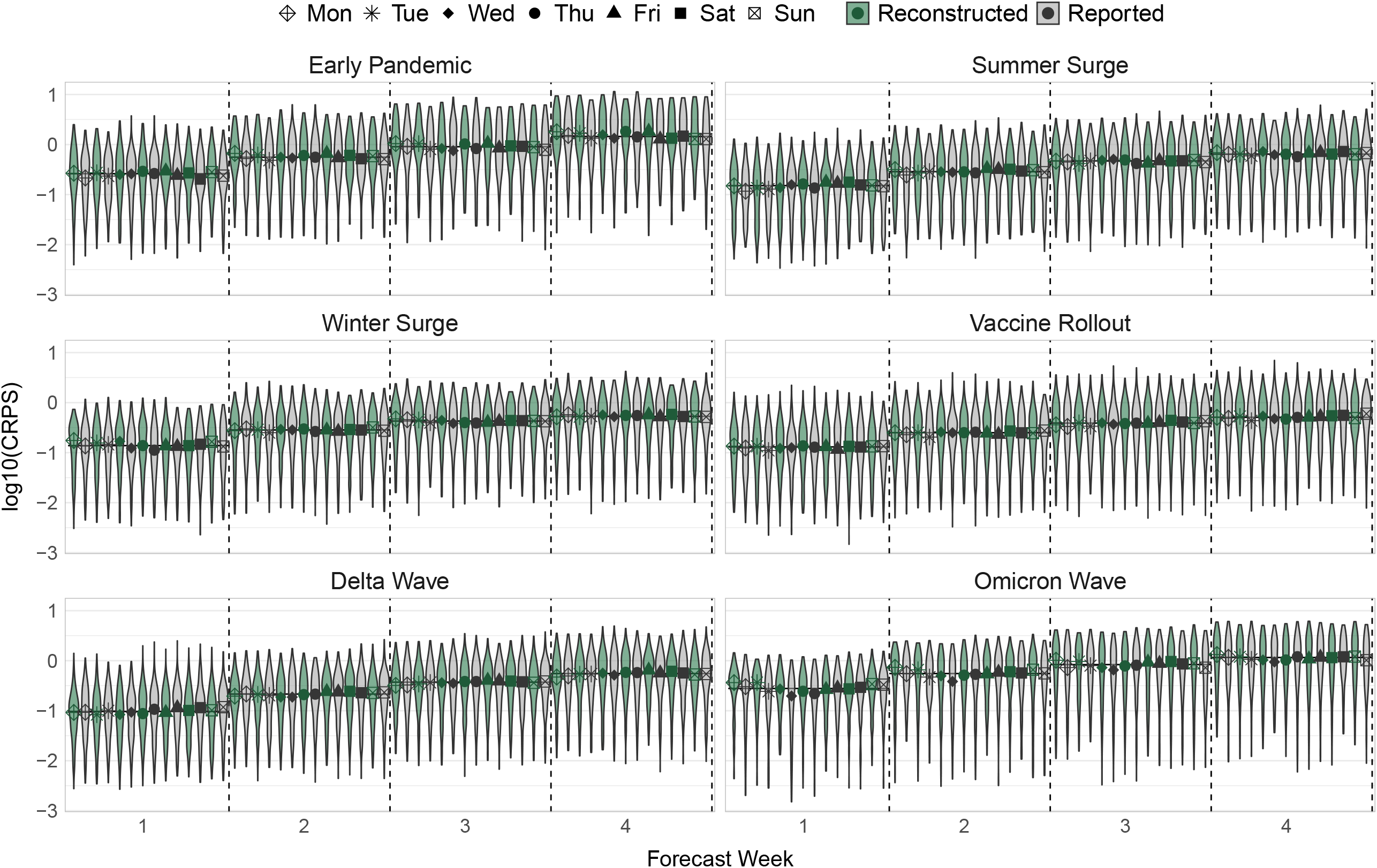
Violin plot showing the distribution of log-transformed (base 10) CRPS values for all included states by outbreak time period, based on the difference in true observed weekly cases and weekly forecasts produced from reconstructed (green) and reported (grey) incidence data. Outbreak phases correspond to those shown in Fig 1. Points denote the median CRPS values, with symbols representing the projection day (last day of past incidence data supplied to the model): Mondays (diamonds with cross) Tuesdays (stars), Wednesdays (solid diamonds), Thursdays (circles), Fridays (triangles), Saturdays (solid squares) and Sundays (squares with cross). The solid black line represents the overall median CRPS score for the forecast week.

## Discussion

The applications of branching process models can be restricted if daily incidence data are unavailable or if there is a transition to aggregated incidence data, such as weekly counts, during prolonged outbreaks [4]. Many US states stopped reporting daily incidence for COVID-19 in summer 2021 [7], and the European Centre for Disease Prevention and Control (ECDC) switched to reporting weekly incidence as early as 14th December 2020 [8]. Notably, branching process models are especially valuable during the early stages of outbreaks when limited data make it challenging to reliably parameterise more complex models. In this study, we applied the EM algorithm to US COVID-19 case incidence data to demonstrate that reconstructed incidence data can achieve comparable forecasting performance to reported incidence data regardless of the projection day (i.e., the last day of past incidence data supplied to the model), US state, or outbreak time period. The findings suggest that while daily reporting may offer some advantages under specific circumstances, such as during the Early Pandemic, the differences in forecasting accuracy are generally minimal. This highlights the viability of reconstructed data as a practical alternative for supporting accurate forecasting efforts in contexts where setting up and maintaining daily reporting is logistically challenging or resource-intensive.

### Forecasting performance across US states

For all included US states combined, model performance was similar regardless of the projection day or whether daily incidence data were reported or reconstructed (Fig 4). Increasing median CRPS values across forecast weeks (indicating worse performance) highlights the inherent uncertainty in predictions with longer forecasting horizons. This pattern is expected, as projections assume that transmissibility remains the same as in the calibration window of past incidence data throughout the forecasting period. Over longer time frames this assumption becomes more unlikely due to factors such as increasing behaviour change and the implementation or relaxation of interventions [1].

When US states were considered independently, there was no clear advantage for the use of either reported or reconstructed incidence data. However, the choice of projection day influenced forecasting performance for some individual states (Fig 4). US states recorded cases by the date they were reported, which is delayed from the actual test date, and were lowest on Sundays and Mondays with a relatively even distribution around a peak on Thursdays (Fig 2), which is similar to the trend observed across all US states by the COVID Tracking Project between March 2020 and February 2021 [34]. This is different to patterns observed in the UK, where case counts were recorded by the date of specimen collection for testing and typically exhibited a ‘weekend-effect’ with lower case counts from Friday to Sunday and relatively stable reporting over the rest of the week [9].

Each US state has its own health department and reporting practices, which lead to state-specific differences in reporting patterns (Fig 2). Most US states reported data with a delay of one or two days, causing a corresponding shift in the ‘weekend-effect’ to the beginning of the following week [34]. There was no obvious correlation between state-specific reporting patterns (Fig 2) and the best performing projection days in forecasts. The model calibration window (see methods) was chosen to smooth out reporting patterns in the data, however this 10-day window would have encompassed either two peaks or two dips in the relative reporting of cases, which could have caused projection day variations in the results. When using reconstructed incidence, calibration windows should always be at least the length of the original data aggregation and longer windows should help smooth out discontinuities in the reconstructed incidence [9].

Given the differences observed in forecasting performance, multiple projection days could be considered for real-time analysis. It is important to also consider the practical utility of forecasts depending on the specific day of the week they are generated. For example, projections made using incidence data up to Sunday are performed on Mondays, which can guide the week’s response [1]. The timing of incidence data publication will also play a role, as some datasets may be updated daily, while others may only be released once a week, limiting flexibility in when forecasts can be performed for real-time response. Therefore, the choice of projection day should balance practical constraints with potential gains in forecasting performance.

### Forecasting performance across outbreak phases

When considering distinct outbreak phases (Fig 1), forecasts made during the Summer Surge to the Delta Wave (June 2020 to November 2021) showed minimal differences in forecasting performance regardless of the dataset and projection day. In contrast, forecasting performance was notably worse in the early phase and the Omicron Wave (Fig 5). Several factors likely contributed to this disparity. Reporting patterns differed most noticeably in the Early Pandemic and Omicron Wave (Fig 3). During the early phase, US states had never collected data at the scale the pandemic demanded before, contending with considerable test shortages and longer reporting delays as testing facilities and health departments adjusted to the initial surge in cases [35]. Despite these limitations, forecasts based on daily reported incidence consistently outperformed those using reconstructed incidence, emphasising the importance of prioritising reported daily incidence for forecasting in the early stages of outbreaks.

During the Omicron Wave, healthcare systems were also under immense strain, which could have led to inconsistencies in reporting and therefore worse forecasting performance. Taking into account different projection days and datasets, there were only small differences between them during the Omicron Wave, but reported incidence data with a Wednesday projection day performed best.

### Limitations

While this study provides valuable insights, there are some limitations. Forecasting performance and the reconstruction of incidence data itself can be influenced by high sensitivity to the specified serial interval (or generation time), as even small errors can lead to significant discrepancies in projected outcomes [36]. Given that the method used does not allow for a time-varying SI, the SI chosen for this analysis aimed to provide a reasonable midpoint for the SI throughout the pandemic. However, this may partly account for the worse performance observed in the Early Pandemic and Omicron Wave when the SI was estimated to be slightly longer and shorter respectively (Fig 5) [27]. It may also explain the improved performance of forecasts using reported data in the early phase. Future sensitivity analyses could explore whether performance improves with varying SI distributions.

Additionally, weekly smoothing occurs when daily incidence data are reconstructed from weekly case counts [9]. This weekly smoothing, combined with the model’s 10-day calibration window, could influence forecasting accuracy during periods of rapid changes in transmission or reporting frequency, such as the Early Pandemic and Omicron Wave. It is also important to consider how the reporting rate varies over the course of an outbreak [37]. For instance, if the healthcare system is overwhelmed and does not have the capacity to handle an influx of cases, the proportion of reported cases would likely be lower than during time periods where the healthcare system is under less pressure. If the proportion of cases reported during the forecast period differs to the calibration window, this will lead to bias in comparisons between forecasts and observed weekly counts. Time-varying reporting rates could be considered in future research.

Although our focus has been on weekly reporting of COVID-19 cases in the US, the approach presented here is broadly applicable to other pathogens and settings. Whilst we have shown that forecasting performance is comparable in this context, determining the optimal reporting frequency for surveillance systems should consider the specific characteristics of the pathogen, as well as the public health infrastructure and context of the country or region of interest.

## Conclusion

Reconstructed incidence data can support reliable and accurate forecasting when daily reported incidence is not available. While daily reported incidence remains critical during the early stages of outbreaks, the findings emphasise the viability and practicality of using reconstructed incidence for diseases where cases are typically reported over longer timescales or when reporting changes arise during prolonged outbreaks. Balancing the benefits of daily reporting with the flexibility offered by reconstructed data can ensure more resilient forecasting methods in real-time outbreak response.

## Data Availability

To reproduce the analysis presented in this study visit https://github.com/mrc-ide/covid19-forecasts-orderly/tree/us_aggregations and run the "us_aggregations_workflow.R" script.

## Acknowledgments

RKN acknowledges funding from the Medical Research Council (MRC) Doctoral Training Partnership (grant reference MR/N014103/1). SB acknowledges funding by The Wellcome Trust (223120/Z/21/Z). JW acknowledges research funding from the Wellcome Trust (grant 102169/Z/13/Z). AC acknowledges the Academy of Medical Sciences Springboard, funded by the Academy of Medical Sciences, Wellcome Trust, the Department for Business, Energy and Industrial Strategy, the British Heart Foundation, and Diabetes UK (reference SBF005 \ 1044). AC also acknowledges funding from the National Institute for Health and Care Research (NIHR) Health Protection Research Unit in Modelling and Health Economics, a partnership between the UK Health Security Agency, Imperial College London and LSHTM (grant code NIHR200908). RKN, SB, JW and AC also acknowledge funding from the MRC Centre for Global Infectious Disease Analysis (reference MR/X020258/1), funded by the UK Medical Research Council (MRC). This UK funded award is carried out in the frame of the Global Health EDCTP3 Joint Undertaking. Disclaimer: The views expressed are those of the author(s) and not necessarily those of the NIHR, UK Health Security Agency or the Department of Health and Social Care. The funders had no role in study design, data collection and analysis, decision to publish, or preparation of the manuscript.

## References

1. Imperial College COVID-19 Response Team. Short-Term Forecasts of COVID-19 Deaths in Multiple Countries;. https://mrc-ide.github.io/covid19-short-term-forecasts/.

2. Chintalapudi N, Battineni G, Sagaro GG, Amenta F. COVID-19 Outbreak Reproduction Number Estimations and Forecasting in Marche, Italy. International Journal of Infectious Diseases. 2020;96:327–333. doi:10.1016/j.ijid.2020.05.029.

3. Bertozzi AL, Franco E, Mohler G, Short MB, Sledge D. The Challenges of Modeling and Forecasting the Spread of COVID-19. Proceedings of the National Academy of Sciences. 2020;117(29):16732–16738. doi:10.1073/pnas.2006520117.

4. Nash RK, Nouvellet P, Cori A. Real-Time Estimation of the Epidemic Reproduction Number: Scoping Review of the Applications and Challenges. PLOS Digital Health. 2022;1(6):e0000052. doi:10.1371/journal.pdig.0000052.

5. COVID-19 Cases WHO COVID-19 Dashboard;. https://data.who.int/dashboards/covid19/cases.

6. Hassan A. Some U.S. States Are Reducing Daily Reporting of Coronavirus Data, Raising Fears of Blind Spots. The New York Times. 2022;.

7. State Data Reporting Reduction Continues Amidst COVID-19 Resurgence;. https://coronavirus.jhu.edu/pandemic-data-initiative/news/state-data-reporting-reduction-continues-amidst-covid-19-resurgence.

8. Download Historical Data (to 14 December 2020) on the Daily Number of New Reported COVID-19 Cases and Deaths Worldwide; 2020. https://www.ecdc.europa.eu/en/publications-data/download-todays-data-geographic-distribution-covid-19-cases-worldwide.

9. Nash RK, Bhatt S, Cori A, Nouvellet P. Estimating the Epidemic Reproduction Number from Temporally Aggregated Incidence Data: A Statistical Modelling Approach and Software Tool. PLOS Computational Biology. 2023;19(8):e1011439. doi:10.1371/journal.pcbi.1011439.

10. Gressani O, Wallinga J, Althaus CL, Hens N, Faes C. EpiLPS: A Fast and Flexible Bayesian Tool for Estimation of the Time-Varying Reproduction Number. PLOS Computational Biology. 2022;18(10):e1010618. doi:10.1371/journal.pcbi.1010618.

11. Holshue ML, DeBolt C, Lindquist S, Lofy KH, Wiesman J, Bruce H, et al. First Case of 2019 Novel Coronavirus in the United States. The New England Journal of Medicine. 2020;382(10):929–936. doi:10.1056/NEJMoa2001191.

12. WHO. Coronavirus Disease (COVID-19) Pandemic;. https://www.who.int/europe/emergencies/situations/covid-19.

13. CDC. CDC Museum COVID-19 Timeline; 2023. https://www.cdc.gov/museum/timeline/covid19.html.

14. State Reporting Frequencies;. https://coronavirus.jhu.edu/data/state-reporting-frequencies.

15. Hale T, Atav T, Hallas L, Kira B, Phillips T, Petherick A, et al. Variation in US States’ Responses to COVID-19. Blavatnik School of Government & Oxford University; 2021. BSG-WP-2020/034.

16. Mehta SH, Clipman SJ, Wesolowski A, Solomon SS. Holiday Gatherings, Mobility and SARS-CoV-2 Transmission: Results from 10 US States Following Thanksgiving. Scientific Reports. 2021;11(1):17328. doi:10.1038/s41598-021-96779-6.

17. Carlos del Rio, Preeti N Malani, Saad B Omer. Confronting the Delta Variant of SARS-CoV-2, Summer 2021. JAMA. 2021;326(11):1001–1002. doi:10.1001/jama.2021.14811.

18. Pryanka Relan, Nkengafac Villyen Motaze, Kavita Kothari, Lisa Askie, Olivier Le Polain de Waroux, Maria D Van Kerkhove, et al. Severity and Outcomes of Omicron Variant of SARS-CoV-2 Compared to Delta Variant and Severity of Omicron Sublineages: A Systematic Review and Metanalysis. BMJ Global Health. 2023;8(7). doi:10.1136/bmjgh-2023-012328.

19. National COVID-19 Preparedness Plan; 2022. https://www.whitehouse.gov/covidplan/.

20. Dong E, Du H, Gardner L. An Interactive Web-Based Dashboard to Track COVID-19 in Real Time. The Lancet Infectious Diseases. 2020;20(5):533–534. doi:10.1016/S1473-3099(20)30120-1.

21. COVID-19 Data Repository by the Center for Systems Science and Engineering (CSSE) at Johns Hopkins University; 2024.

22. U.S. Census Bureau QuickFacts: United States;. https://www.census.gov/quickfacts/fact/table/.

23. Mrc-Ide/EpiEstim; 2024. MRC Centre for Global Infectious Disease Analysis.

24. Nouvellet P, Cori A, Garske T, Blake IM, Dorigatti I, Hinsley W, et al. A Simple Approach to Measure Transmissibility and Forecast Incidence. Epidemics. 2018;22:29–35. doi:10.1016/j.epidem.2017.02.012.

25. Cori A, Ferguson NM, Fraser C, Cauchemez S. A New Framework and Software to Estimate Time-Varying Reproduction Numbers during Epidemics. American Journal of Epidemiology. 2013;178(9):1505–1512. doi:10.1093/aje/kwt133.

26. Fraser C. Estimating Individual and Household Reproduction Numbers in an Emerging Epidemic. PLOS ONE. 2007;2(8):e758. doi:10.1371/journal.pone.0000758.

27. Madewell ZJ, Yang Y, Longini IM, Halloran ME, Vespignani A, Dean NE. Rapid Review and Meta-Analysis of Serial Intervals for SARS-CoV-2 Delta and Omicron Variants. BMC infectious diseases. 2023;23(1):429. doi:10.1186/s12879-023-08407-5.

28. Wang Y, Teunis P. Strongly Heterogeneous Transmission of COVID-19 in Mainland China: Local and Regional Variation. Frontiers in Medicine. 2020;7. doi:10.3389/fmed.2020.00329.

29. Bhatia S, Parag KV, Wardle J, Nash RK, Imai N, Elsland SLV, et al. Retrospective Evaluation of Real-Time Estimates of Global COVID-19 Transmission Trends and Mortality Forecasts. PloS One. 2023;18(10):e0286199. doi:10.1371/journal.pone.0286199.

30. Sangeeta Bhatia, Robert Ashton. Mrc-Ide/Jointlyr; 2022. MRC Centre for Global Infectious Disease Analysis.

31. Bosse NI, Abbott S, Cori A, van Leeuwen E, Bracher J, Funk S. Scoring Epidemiological Forecasts on Transformed Scales. PLOS Computational Biology. 2023;19(8):e1011393. doi:10.1371/journal.pcbi.1011393.

32. Bosse NI, Gruson H, Cori A, van Leeuwen E, Funk S, Abbott S. Evaluating Forecasts with Scoringutils in R. arXiv. 2022;doi:10.48550/ARXIV.2205.07090.

33. Jordan A, Krüger F, Lerch S. Evaluating Probabilistic Forecasts with scoringRules. Journal of Statistical Software. 2019;90(12):1–37. doi:10.18637/jss.v090.i12.

34. Hannah Hoffman. Analysis & Updates How Day-of-Week Effects Impact COVID-19 Data;. https://covidtracking.com/analysis-updates/how-day-of-week-effects-impact-covid-19-data.

35. Schneider EC. Failing the Test — The Tragic Data Gap Undermining the U.S. Pandemic Response. New England Journal of Medicine. 2020;383(4):299–302. doi:10.1056/NEJMp2014836.

36. Gostic KM, McGough L, Baskerville EB, Abbott S, Joshi K, Tedijanto C, et al. Practical Considerations for Measuring the Effective Reproductive Number, Rt. PLOS Computational Biology. 2020;16(12):e1008409. doi:10.1371/journal.pcbi.1008409.

37. Spannaus A, Papamarkou T, Erwin S, Christian JB. Inferring the Spread of COVID-19: The Role of Time-Varying Reporting Rate in Epidemiological Modelling. Scientific Reports. 2022;12(1):10761. doi:10.1038/s41598-022-14979-0.

